# Causal associations between salt intake with body mass, shape and composition: A two-sample Mendelian randomization study

**DOI:** 10.1101/2020.05.01.20087007

**Authors:** Qi Feng, Shuai Yuan, Qian Yang, Yingchang Lu, Ruth J.F. Loos, Gloria H.Y. Li, Yue Fei, Man Fung Tsoi, Ching Lung Cheung, Bernard M.Y. Cheung

## Abstract

**Background:** Observational studies have found associations between salt intake with obesity, body shape and composition; but the findings may be biased by residual confounding.

**Objective:** To analyze the causal associations between salt intake and obesity measures in both sex-combined and sex-specific models.

**Designs:** This was a two-sample Mendelian randomization study. Genome-wide association (GWA) studies of urinary sodium secretion (UNa, a surrogate for salt intake), body mass index (BMI), BMI-adjusted waist-to-hip ratio (WHR), body fat (BF) percentage and estimated glomerular filtration rate (eGFR) were identified. We initially extracted fifty SNPs associated with UNa at GWA significance level of 5 × 10^−8^, but further removed those SNPs with potential horizontal pleiotropy. Univariable MR and multivariable MR with adjustment for eGFR were performed. Inverse-variance weighted MR was performed as the primary analysis, with MR-Egger methods as sensitivity analysis. The potential bidirectional association between BMI and UNa was investigated. All exposure and outcomes were continuous, and the effect measure was linear regression coefficients (beta) and their 95% confidence intervals (95%CI).

**Results:** UNa was causally associated with increased BMI in both men (eGFR-adjusted beta 0.443 (0.163 to 0.724)) and women (0.594 (0.333 to 0.855)). UNa caused BF percentage increase in men (0.622 (0.268 to 0.976)) and women (0.334 (0.007 to 0.662)). UNa significantly elevated BMI-adjusted WHR in men (0.321 (0.094 to 0.548)), but not in women (0.170 (−0.052 to 0.391)). Additionally, we found that BMI causally increased UNa (0.043 (0.023 to 0.063)).

**Conclusions:** Salt intake increased BMI and BF percentage. Salt intake affects male body shape by increasing BMI-adjusted WHR, but showed no effects on female body shape. The bidirectional association between BMI and UNa suggested that salt reduction measures and weight reduction measures should be implemented simultaneously to break the vicious cycle and gain more health benefits.

## Introduction

Obesity has been a global public health concern with increasingly substantial disease burdens (1–3). Obesity is believed to result from a complex relationship among psychosocial, biological and behavioral factors, such as genetic susceptibility, excessive calorie intake and physical inactivity (4). Other factors also contribute independently to development of obesity. (4,5) Obesity, commonly measured with body mass index (BMI, body weight in kilogram/squared height in meter), is a well-recognized risk factor for a range of health conditions and diseases, including cardiovascular diseases (6), diabetes (7,8) and cancers (9–11). BMI is easy to measure and calculate; nevertheless, body shape and body composition are sometimes stronger predictors for health outcomes than BMI (12,13).

Dietary salt intake is known to be associated with disease conditions, such as blood pressure (14), cardiovascular diseases (15), and diabetic complications (16). Salt intake can be measured with food frequency questionnaire or estimated from urinary sodium secretion (UNa), the latter of which is considered free of recall bias and more objective (17). Observational studies have suggested its effect on obesity, independent of total energy intake and fluid consumption. A systematic review published in 2015 (18) found that both dietary salt intake and UNa were positively associated with BMI, although with substantial heterogeneity. A recent cross-sectional study (19) showed that this positive association was directionally concordant across Asian and Western populations, with varying effect sizes. Another cross-sectional study (20) revealed an effect modified by sex, because the effect was observed only in women but not men. Salt intake was also demonstrated to be correlated with body shape and body composition measures, such as waist circumference (WC) (17,21), waist-to-hip ratio (WHR, waist circumference/hip circumference) (17), body fat (BF) mass and lean mass (20).

However, so far, the majority of the evidence has been generated from observational studies, mainly cross-sectional studies, in which residual confounding and reverse causation are likely. So far, few randomized trials on salt reduction have reported obesity-related outcomes (22,23). Mendelian randomization (MR), by using single nucleotide polymorphisms (SNPs) as instrumental variables to investigate the causal link between two phenotypes, is helpful in causal inference (24). The objective of this study is to examine the causal association between UNa with body mass, shape and composition using two-sample MR approach.

## Materials and Methods

In this study, we used summary-level statistics from relevant GWA studies, where ethical approval and patient consent had been obtained. The exposure of interest was UNa, a commonly used surrogate marker for salt intake (25). The outcomes included BMI, body shape and body composition. For body shape, we used BMI-adjusted WHR as the primary outcome, while WC, hip circumference (HC), WHR, BMI-adjusted WC, BMI-adjusted HC were secondary outcomes. For body composition, we used BF percentage as the primary outcome, while whole-body lean mass (WLM) and appendicular lean mass (ALM) as secondary outcomes. Since sex has been suggested as a potential effect modifier in obesity-related outcomes (26–28), we investigated sex-specific effects in men and women separately for the outcomes, if possible.

## SNP selection

A GWA study of 446,237 European-ancestry individuals from UK Biobank (29) identified 50 lead SNPs associated with UNa that reached significance level of 5 × 10^−8^ (Supplementary **table** 1). Spot urine sample collection and storage in UK Biobank have been described elsewhere (30). UNa concentration was measured by the ion-selective electrode method. The effect between genetic variants and log-transformed UNa concentration was adjusted for age, sex, array information and ancestral principal components. The SNPs are located in different gene regions and distributed independently (not in linkage disequilibrium), and explain 6.4% of total variance of the trait (29). If an SNP could not be matched in the GWA studies of an outcome, a proxy in linkage disequilibrium with the SNP (R^2^ > 0.80) would be identified; if no proper proxy was identified, the unmatched SNP would be removed from analysis. Relevant traits associated with the SNPs were also searched in the PhenoScanner v2 database at a significance level of 5 × 10^−8^ for potential pleiotropy.

## Outcome data

We extracted relevant summary-level data of the outcomes from GWA studies of BMI (31), body shape (32) and body composition (33,34). The BMI GWA study (31) included 322,154 European-ancestry individuals (152,882 men and 171,963 women) from 125 cohorts. The effect was adjusted for age, squared age, ancestral principal components and study-specific covariates. The body shape GWA study (32) included 211,088 European-ancestry individuals (93,480 men and 116,742 women), and provided data for BMI-adjusted and unadjusted WC, HC, and WHR, additionally adjusted for age, squared age and ancestral principal components. The BF percentage GWA study (34) included 89,297 European-ancestry individuals (44,429 men and 45,525 women), and was adjusted for age, squared age and ancestral principal components. The GWA study for WLM and ALM (33) included 38,292 and 28,330 European-ancestry individuals, respectively, and was adjusted for age, sex, squared age, height, body fat percentage and ancestral principal components. Body composition (BF, WLM and ALM) was measured with either bioimpedance analysis or dual-energy X-ray absorptiometry (32,33). Sex-combined and sex-specific outcome data were available from the GWA studies of BMI, body shape and BF percentage. The GWA study for WLM and ALM provided sex-combined data only.

## Covariate

Renal function is associated with both UNa secretion (35) and obesity (36,37), which is a potential confounder in the associations of interest. We identified a GWA study (38) of estimated glomerular filtration rate (eGFR), which included 567,460 European-ancestry individuals. Linear regression of log-transformed eGFR was adjusted for age, sex and ancestry principal components.

## Statistical analysis

Previous studies (29) have suggested that some of the UNa-associated SNPs are related to adiposity phenotypes, which could introduce horizontal pleiotropy and violate one of the underlying assumptions of MR. To minimize potential pleiotropy, we further removed SNPs that: (1) were associated with the outcomes of interest at significance level of 5 × 10^−8^, or (2) were identified as outliers by the MR Pleiotropy RESidual Sum and Outlier (MR-PRESSO) outlier tests (39). Therefore, the included SNPs for each outcome may differ but remained a subset of the 50 SNPs identified by the UNa GWA study (29). Supplementary file 1 shows the characteristics of the SNPs included in the MR analysis for each outcome. F statistic was used to measure the strength of instrumental variables, and a F statistic > 10 suggests strong instrument variables. (40)

All GWA summary-level data were harmonized with the method of Hartwig *et al*. (41), so that the effect estimates of SNPs on exposure, outcome and covariate were shown for the same alleles. Univariable MR analyses were performed with inverse-variance weighted method as primary analysis, while weighted median method and MR-Egger regression method as sensitivity analysis. Multivariable MR analyses adjusted for eGFR with inverse-variance weighted method were performed as primary analysis, while MR-Egger method as secondary analysis. The intercept test for MR-Egger regression was used to examine potential residual horizontal pleiotropy, with a P value < 0.05 suggesting pleiotropy.

To investigate sex-specific effects, we extracted male-specific and female-specific outcome data from the GWA studies, where applicable (31,32,34), and performed MR analyses accordingly. Sex-specific effects on WLM and ALM were not estimated, because the data were unavailable. To examine the effect difference between men and women, we performed a heterogeneity test similar to that in meta-analysis, using Cochrane’s Q test. A P-value for Cochrane’s Q test < 0.10 indicates statistical significance.

To investigate potential bidirectional association between UNa and BMI, we performed MR analysis with BMI as the exposure and UNa as the outcome. Ninety-seven BMI-associated SNPs were extracted from the BMI GWA study (31), but 84 SNPs were finally included after removing those unmatched with UNa GWA data (29) and/or identified as outliers by the MR-PRESSO outlier tests. Similarly, univariable and multivariable MR analyses were performed.

A second way to deal with potential confounding effect of eGFR, alternative to the multivariable MR, was to remove the SNPs associated with eGFR. We found that rs1260326 (located in *GCKR* gene) was associated with eGFR at a GWA significance level of 5 × 10^−8^ in Phenoscanner v2 database; therefore, a sensitivity analysis by removing rs1260326 in univariable MR was performed as sensitivity analysis. Additional sensitivity analyses were performed by including outlier SNPs detected by the MR-PRESSO outlier tests. Since all outcomes were continuous, the effect measure was linear regression coefficient beta with its 95% confidence interval (CI), which should be interpreted as the change in units of standard deviation (SD) in the outcome when the log-UNa level increases 1 SD. Statistical significance was indicated by p value < 0.05. We did not apply Bonferroni correction, although we employed three primary outcomes, because their effects were estimated from three independent samples of individuals, instead of one. All statistical analysis was conducted with “*MendelianRandomization*” (version 0.4.1) and “*MRPRESSO*” (version 1.0) packages in R environment. **Supplementary figure 1** shows the flowchart of data selection and analysis.

We planned to perform power calculations based on the method of Brion *et al*. (42), which required the beta estimates from our study and another beta estimates from observational analysis. However, the UNa GWA study analyzed log-UNa concentration (in log-mmol/L), while previous observational studies analyzed UNa concentration in a natural unit (mmol/L) or converted it to 24-hour UNa (mmol/d or g/d) or daily salt intake (g/d), thus making it impossible to make direct comparisons of the two beta estimates in the absence of individual participant data.

## Results

The proportions of exposure variance explained by the instrument variables ranged from 4.036% to 5.565% (mean 4.780%), and the according F statistics ranged from 536.146 to 613.340 (mean 587.078), suggesting strong instrumental variables. (**Table** 1) Supplementary file 1 showed detailed information on the SNPs included in the MR analysis for each outcome.

**Table 1.**
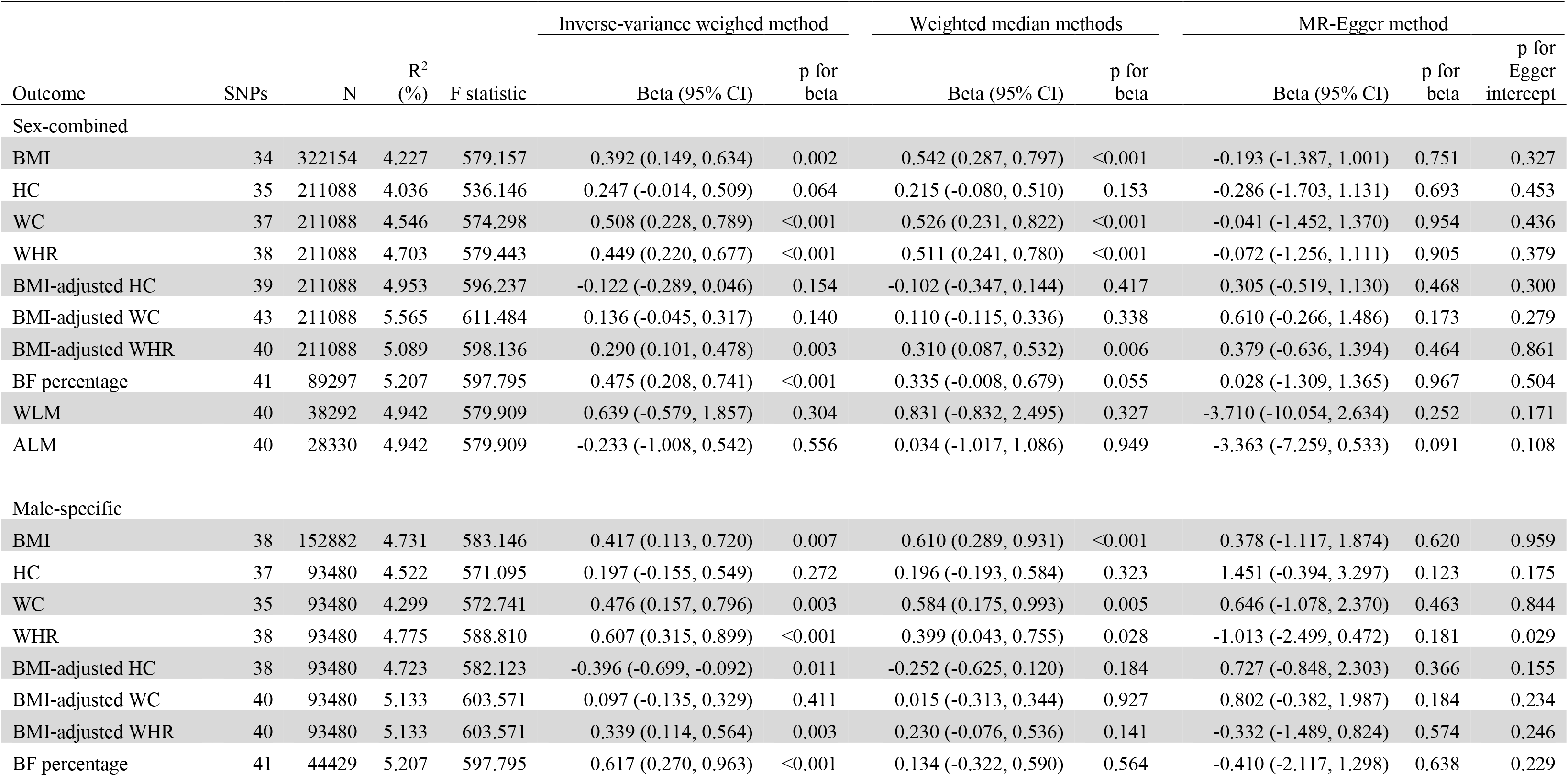
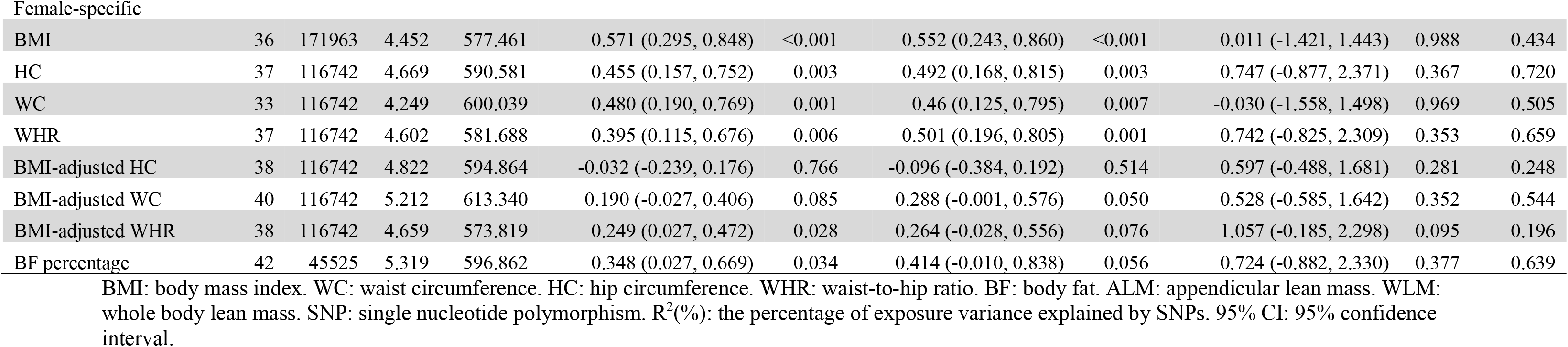
Results of univariable Mendelian randomization analyses of sex-combined and sex-specific association between urinary sodium secretion with body mass, shape and composition outcomes.

| Outcome | SNPs | N | R<sup>2</sup><br>(%) | F statistic | Inverse-variance weighed method |  | Weighted median methods |  | MR-Egger method |  |  |
| --- | --- | --- | --- | --- | --- | --- | --- | --- | --- | --- | --- |
|  |  |  |  |  | Beta (95% CI) | p for<br>beta | Beta (95% CI) | p for<br>beta | Beta (95% CI) | p for<br>beta | p for<br>Egger<br>intercept |
| Sex-combined |  |  |  |  |  |  |  |  |  |  |  |
| BMI | 34 | 322154 | 4.227 | 579.157 | 0.392 (0.149, 0.634) | 0.002 | 0.542 (0.287, 0.797) | <0.001 | -0.193 (-1.387, 1.001) | 0.751 | 0.327 |
| HC | 35 | 211088 | 4.036 | 536.146 | 0.247 (-0.014, 0.509) | 0.064 | 0.215 (-0.080, 0.510) | 0.153 | -0.286 (-1.703, 1.131) | 0.693 | 0.453 |
| WC | 37 | 211088 | 4.546 | 574.298 | 0.508 (0.228, 0.789) | <0.001 | 0.526 (0.231, 0.822) | <0.001 | -0.041 (-1.452, 1.370) | 0.954 | 0.436 |
| WHR | 38 | 211088 | 4.703 | 579.443 | 0.449 (0.220, 0.677) | <0.001 | 0.511 (0.241, 0.780) | <0.001 | -0.072 (-1.256, 1.111) | 0.905 | 0.379 |
| BMI-adjusted HC | 39 | 211088 | 4.953 | 596.237 | -0.122 (-0.289, 0.046) | 0.154 | -0.102 (-0.347, 0.144) | 0.417 | 0.305 (-0.519, 1.130) | 0.468 | 0.300 |
| BMI-adjusted WC | 43 | 211088 | 5.565 | 611.484 | 0.136 (-0.045, 0.317) | 0.140 | 0.110 (-0.115, 0.336) | 0.338 | 0.610 (-0.266, 1.486) | 0.173 | 0.279 |
| BMI-adjusted WHR | 40 | 211088 | 5.089 | 598.136 | 0.290 (0.101, 0.478) | 0.003 | 0.310 (0.087, 0.532) | 0.006 | 0.379 (-0.636, 1.394) | 0.464 | 0.861 |
| BF percentage | 41 | 89297 | 5.207 | 597.795 | 0.475 (0.208, 0.741) | <0.001 | 0.335 (-0.008, 0.679) | 0.055 | 0.028 (-1.309, 1.365) | 0.967 | 0.504 |
| WLM | 40 | 38292 | 4.942 | 579.909 | 0.639 (-0.579, 1.857) | 0.304 | 0.831 (-0.832, 2.495) | 0.327 | -3.710 (-10.054, 2.634) | 0.252 | 0.171 |
| ALM | 40 | 28330 | 4.942 | 579.909 | -0.233 (-1.008, 0.542) | 0.556 | 0.034 (-1.017, 1.086) | 0.949 | -3.363 (-7.259, 0.533) | 0.091 | 0.108 |
| Male-specific |  |  |  |  |  |  |  |  |  |  |  |
| BMI | 38 | 152882 | 4.731 | 583.146 | 0.417 (0.113, 0.720) | 0.007 | 0.610 (0.289, 0.931) | <0.001 | 0.378 (-1.117, 1.874) | 0.620 | 0.959 |
| HC | 37 | 93480 | 4.522 | 571.095 | 0.197 (-0.155, 0.549) | 0.272 | 0.196 (-0.193, 0.584) | 0.323 | 1.451 (-0.394, 3.297) | 0.123 | 0.175 |
| WC | 35 | 93480 | 4.299 | 572.741 | 0.476 (0.157, 0.796) | 0.003 | 0.584 (0.175, 0.993) | 0.005 | 0.646 (-1.078, 2.370) | 0.463 | 0.844 |
| WHR | 38 | 93480 | 4.775 | 588.810 | 0.607 (0.315, 0.899) | <0.001 | 0.399 (0.043, 0.755) | 0.028 | -1.013 (-2.499, 0.472) | 0.181 | 0.029 |
| BMI-adjusted HC | 38 | 93480 | 4.723 | 582.123 | -0.396 (-0.699, -0.092) | 0.011 | -0.252 (-0.625, 0.120) | 0.184 | 0.727 (-0.848, 2.303) | 0.366 | 0.155 |
| BMI-adjusted WC | 40 | 93480 | 5.133 | 603.571 | 0.097 (-0.135, 0.329) | 0.411 | 0.015 (-0.313, 0.344) | 0.927 | 0.802 (-0.382, 1.987) | 0.184 | 0.234 |
| BMI-adjusted WHR | 40 | 93480 | 5.133 | 603.571 | 0.339 (0.114, 0.564) | 0.003 | 0.230 (-0.076, 0.536) | 0.141 | -0.332 (-1.489, 0.824) | 0.574 | 0.246 |
| BF percentage | 41 | 44429 | 5.207 | 597.795 | 0.617 (0.270, 0.963) | <0.001 | 0.134 (-0.322, 0.590) | 0.564 | -0.410 (-2.117, 1.298) | 0.638 | 0.229 |
| Female-specific |  |  |  |  |  |  |  |  |  |  |  |
| BMI | 36 | 171963 | 4.452 | 577.461 | 0.571 (0.295, 0.848) | <0.001 | 0.552 (0.243, 0.860) | <0.001 | 0.011 (-1.421, 1.443) | 0.988 | 0.434 |
| HC | 37 | 116742 | 4.669 | 590.581 | 0.455 (0.157, 0.752) | 0.003 | 0.492 (0.168, 0.815) | 0.003 | 0.747 (-0.877, 2.371) | 0.367 | 0.720 |
| WC | 33 | 116742 | 4.249 | 600.039 | 0.480 (0.190, 0.769) | 0.001 | 0.46 (0.125, 0.795) | 0.007 | -0.030 (-1.558, 1.498) | 0.969 | 0.505 |
| WHR | 37 | 116742 | 4.602 | 581.688 | 0.395 (0.115, 0.676) | 0.006 | 0.501 (0.196, 0.805) | 0.001 | 0.742 (-0.825, 2.309) | 0.353 | 0.659 |
| BMI-adjusted HC | 38 | 116742 | 4.822 | 594.864 | -0.032 (-0.239, 0.176) | 0.766 | -0.096 (-0.384, 0.192) | 0.514 | 0.597 (-0.488, 1.681) | 0.281 | 0.248 |
| BMI-adjusted WC | 40 | 116742 | 5.212 | 613.340 | 0.190 (-0.027, 0.406) | 0.085 | 0.288 (-0.001, 0.576) | 0.050 | 0.528 (-0.585, 1.642) | 0.352 | 0.544 |
| BMI-adjusted WHR | 38 | 116742 | 4.659 | 573.819 | 0.249 (0.027, 0.472) | 0.028 | 0.264 (-0.028, 0.556) | 0.076 | 1.057 (-0.185, 2.298) | 0.095 | 0.196 |
| BF percentage | 42 | 45525 | 5.319 | 596.862 | 0.348 (0.027, 0.669) | 0.034 | 0.414 (-0.010, 0.838) | 0.056 | 0.724 (-0.882, 2.330) | 0.377 | 0.639 |

BMI: body mass index. WC: waist circumference. HC: hip circumference. WHR: waist-to-hip ratio. BF: body fat. ALM: appendicular lean mass. WLM: whole body lean mass. SNP: single nucleotide polymorphism. $R^2(\%)$ : the percentage of exposure variance explained by SNPs. 95% CI: 95% confidence interval.

## Univariable MR

In sex-combined analyses, MR analyses with inverse-variance weighted method showed positive causal associations between UNa with BMI (beta (95%CI): 0.392 (0.149 to 0.634); **Figure 1**(A)), BMI-adjusted WHR (0.290 (0.101 to 0.478); **Figure 2**(A)) and BF percentage (0.475 (0.208 to 0.741); **Figure 3**(A)). UNa was not associated with BMI-adjusted WC, BMI-adjusted HC, WLM or ALM. MR-Egger intercept tests revealed no horizontal pleiotropy. (Table 1)

**Figure 1:**
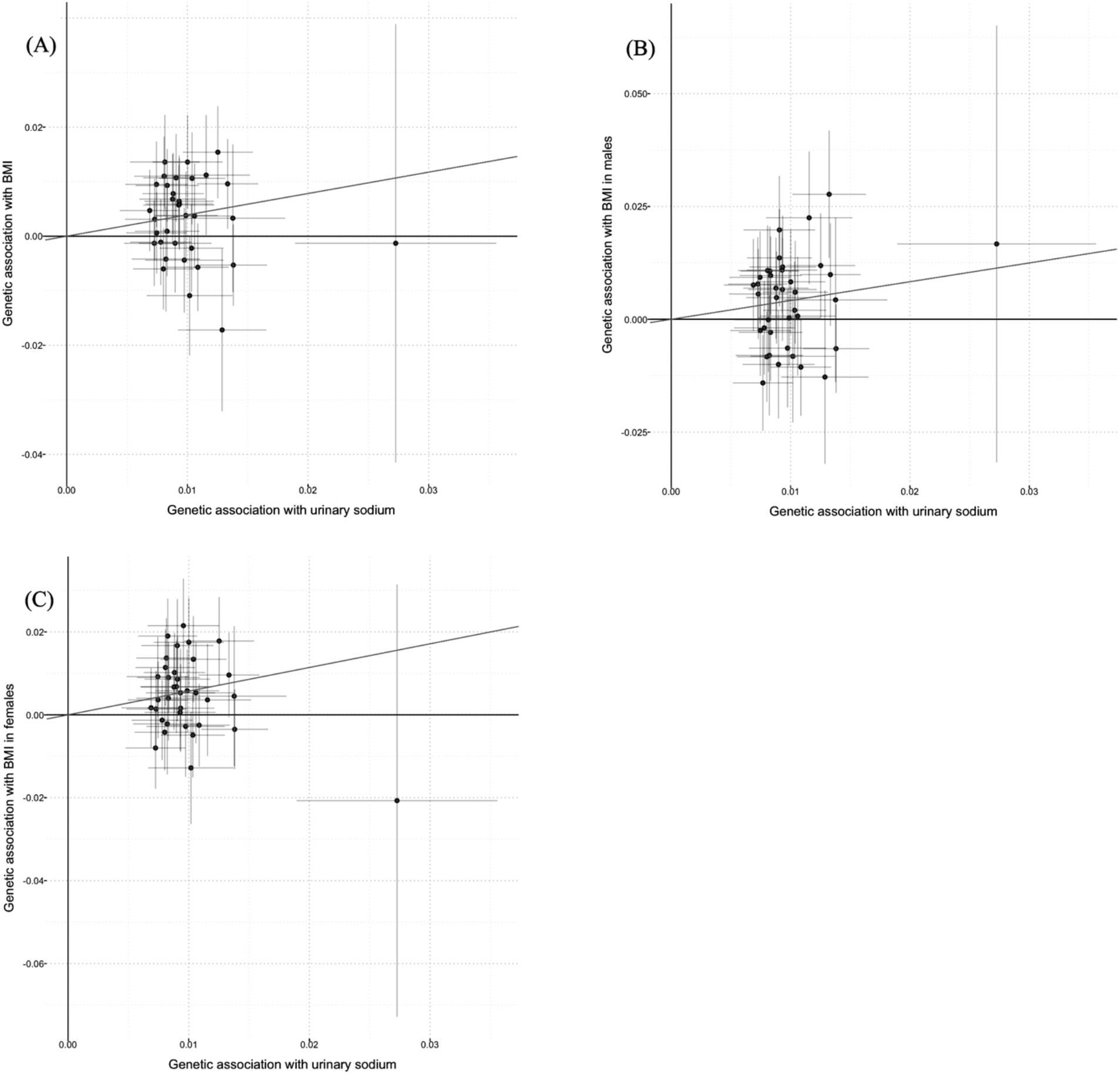
Scatter plots for Mendelian randomization analysis of urinary sodium and BMI. (A) sex-combined, (B) male-specific, (C) female-specific. Horizontal axis: SNPs’ association with urinary sodium. Vertical axis: SNPs’ association with BMI. The fitted line showed the results of the inverse-variance weighted method in univariable MR.

**Figure 2:**
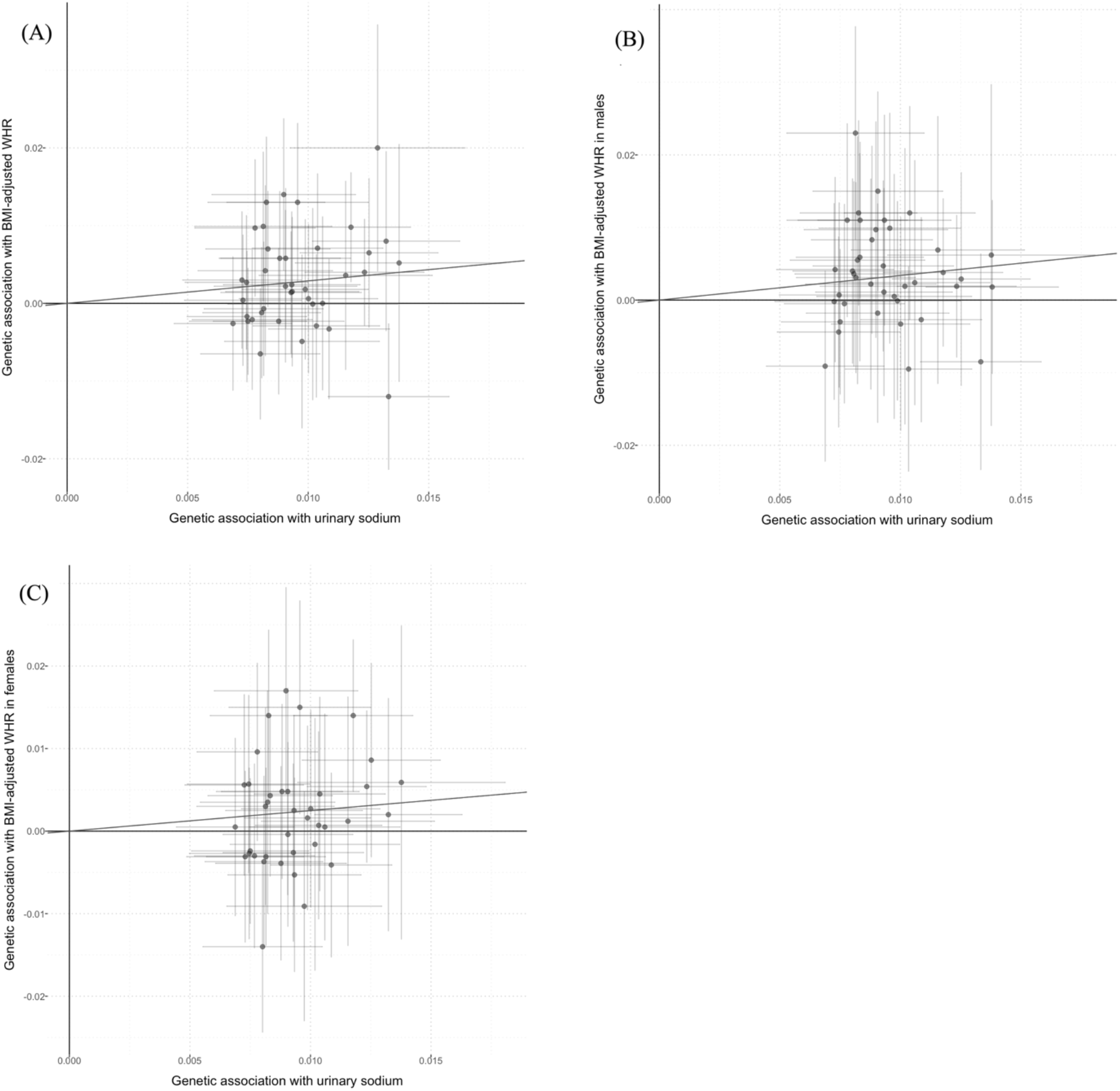
Scatter plots for Mendelian randomization analysis of urinary sodium and BMI-adjusted waist-to-hip ratio. (A) sex-combined, (B) male-specific, (C) female-specific. Horizontal axis: SNPs’ association with urinary sodium. Vertical axis: SNPs’ association with BMI-adjusted waist-to-hip ratio. The fitted line showed the results of the inverse-variance weighted method in univariable MR.

**Figure 3:**
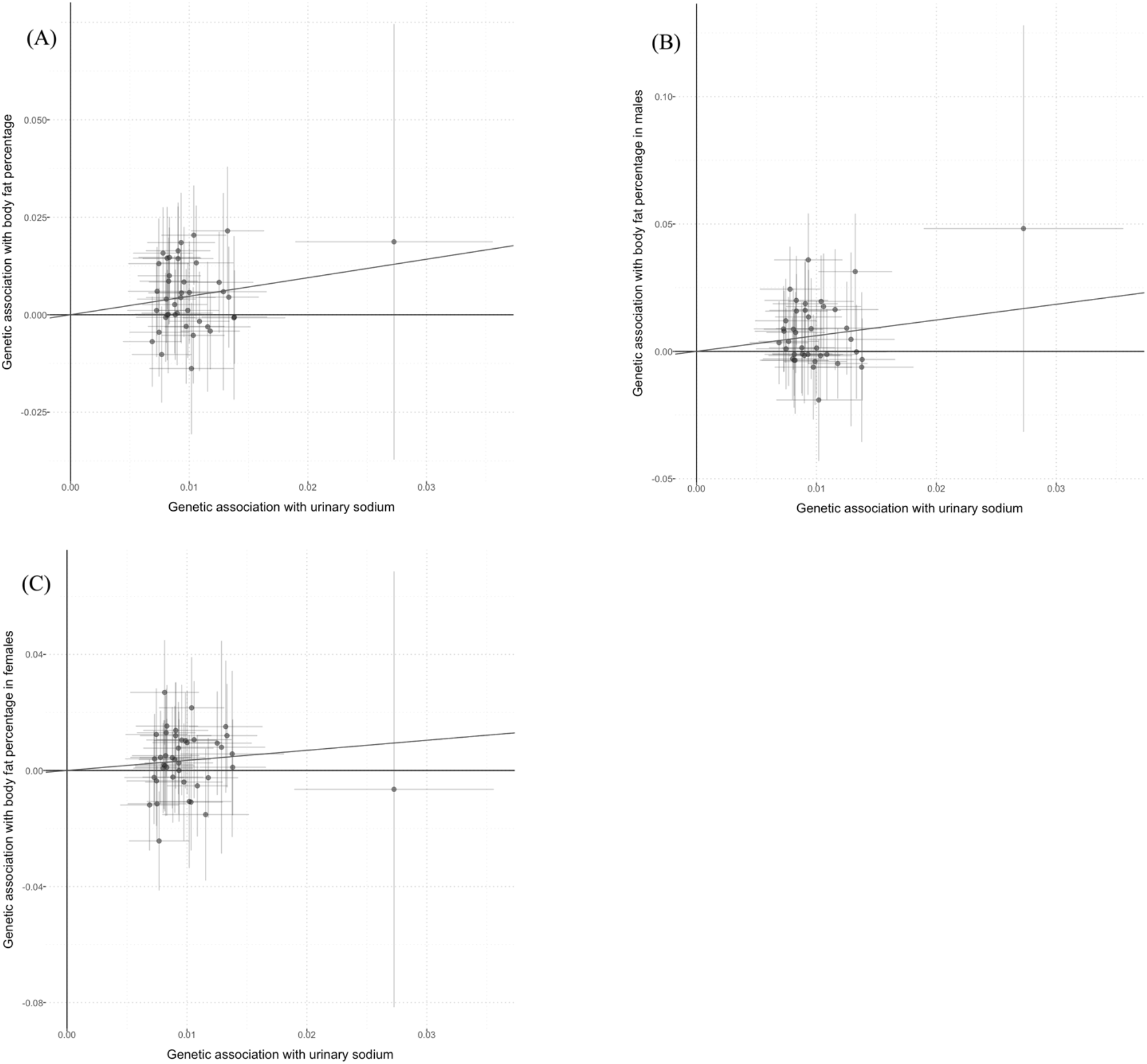
Scatter plots for Mendelian randomization analysis of urinary sodium and body fat percentage. (A) sex-combined, (B) male-specific, (C) female-specific. Horizontal axis: SNPs’ association with urinary sodium. Vertical axis: SNPs’ association with body fat percentage. The fitted line showed the results of the inverse-variance weighted method in univariable MR.

BMI was positively related to UNa with the effect size of 0.042 (0.022 to 0.062), consistent with the weighted median estimator (0.042 (0.019 to 0.066)). MR-Egger intercept test showed no pleiotropy (p = 0.119). This suggested a bidirectional association between UNa and BMI.

In men, UNa was positively associated with BMI (0.417 (0.113 to 0.720); Figure 1(B)), BMI-adjusted WHR (0.339 (0.114 to 0.564); Figure 2(B)) and BF percentage (0.617 (0.270 to 0.963); Figure 3(B)), but negatively with BMI-adjusted HC (−0.396 (−0.699 to −0.092)). No pleiotropy was observed according to MR-Egger intercept test, except WHR (p = 0.029), which was a secondary outcome. UNa was associated with WC but not BMI-adjusted WC, which suggested that BMI might mediate the association between UNa with WC. (Table 2)

**Table 2:**
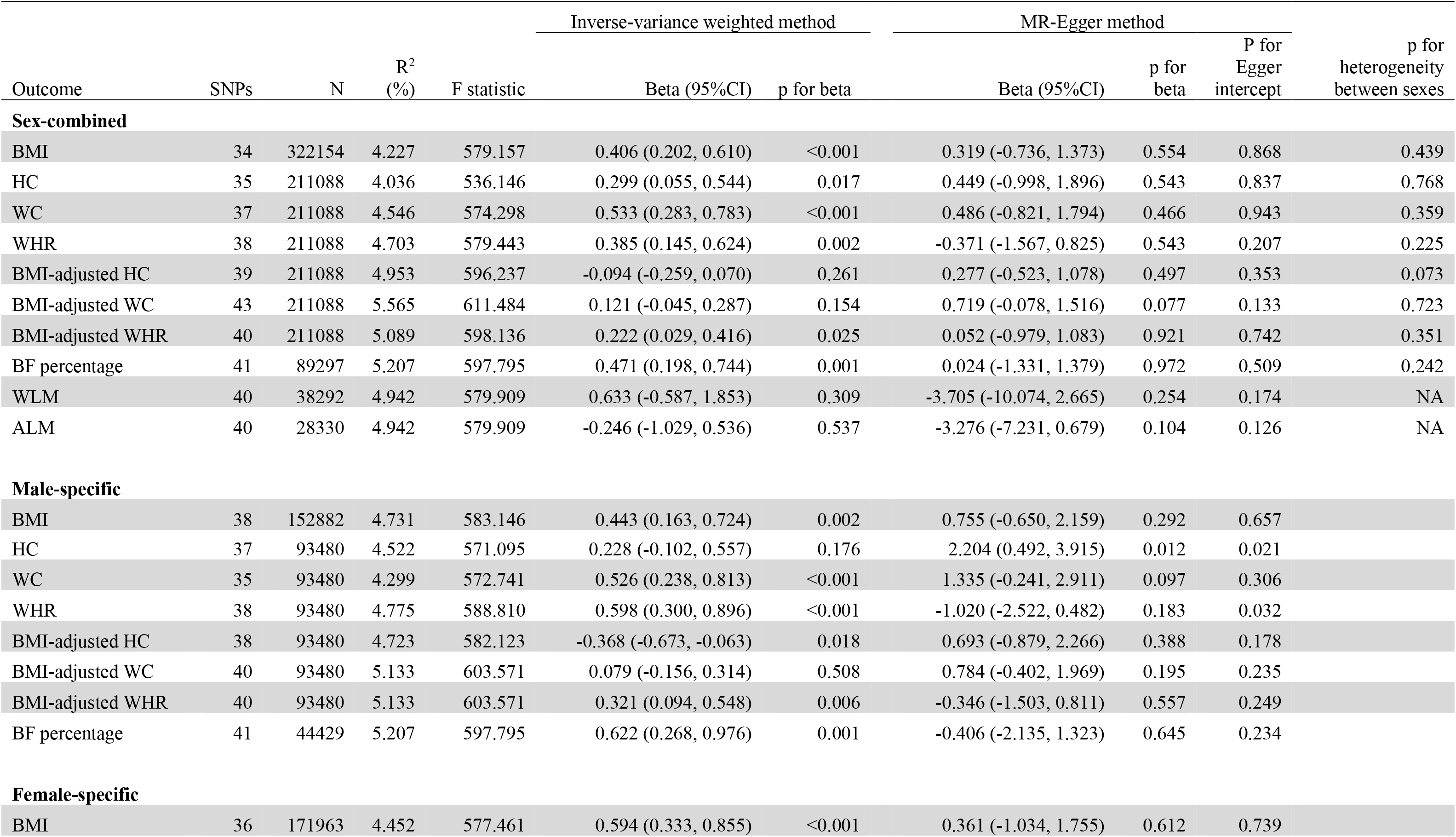
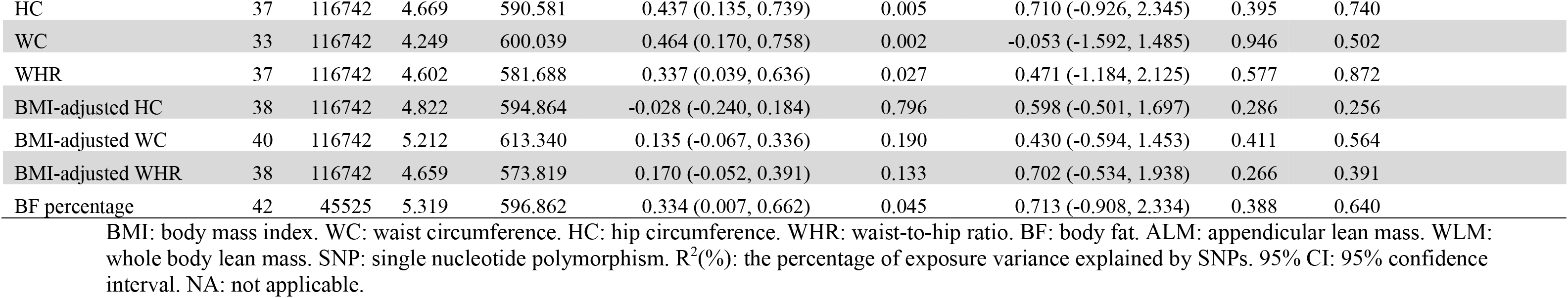
Results of multivariable Mendelian randomization analysis (inverse-variance weighted method) adjusted for estimated glomerular filtration rate

| Outcome | SNPs | N | R<sup>2</sup><br>(%) | F statistic | Inverse-variance weighted method |  | MR-Egger method |  |  | p for<br>heterogeneity<br>between sexes |
| --- | --- | --- | --- | --- | --- | --- | --- | --- | --- | --- |
|  |  |  |  |  | Beta (95%CI) | p for beta | Beta (95%CI) | p for<br>beta | P for<br>Egger<br>intercept |  |
| Sex-combined |  |  |  |  |  |  |  |  |  |  |
| BMI | 34 | 322154 | 4.227 | 579.157 | 0.406 (0.202, 0.610) | <0.001 | 0.319 (-0.736, 1.373) | 0.554 | 0.868 | 0.439 |
| HC | 35 | 211088 | 4.036 | 536.146 | 0.299 (0.055, 0.544) | 0.017 | 0.449 (-0.998, 1.896) | 0.543 | 0.837 | 0.768 |
| WC | 37 | 211088 | 4.546 | 574.298 | 0.533 (0.283, 0.783) | <0.001 | 0.486 (-0.821, 1.794) | 0.466 | 0.943 | 0.359 |
| WHR | 38 | 211088 | 4.703 | 579.443 | 0.385 (0.145, 0.624) | 0.002 | -0.371 (-1.567, 0.825) | 0.543 | 0.207 | 0.225 |
| BMI-adjusted HC | 39 | 211088 | 4.953 | 596.237 | -0.094 (-0.259, 0.070) | 0.261 | 0.277 (-0.523, 1.078) | 0.497 | 0.353 | 0.073 |
| BMI-adjusted WC | 43 | 211088 | 5.565 | 611.484 | 0.121 (-0.045, 0.287) | 0.154 | 0.719 (-0.078, 1.516) | 0.077 | 0.133 | 0.723 |
| BMI-adjusted WHR | 40 | 211088 | 5.089 | 598.136 | 0.222 (0.029, 0.416) | 0.025 | 0.052 (-0.979, 1.083) | 0.921 | 0.742 | 0.351 |
| BF percentage | 41 | 89297 | 5.207 | 597.795 | 0.471 (0.198, 0.744) | 0.001 | 0.024 (-1.331, 1.379) | 0.972 | 0.509 | 0.242 |
| WLM | 40 | 38292 | 4.942 | 579.909 | 0.633 (-0.587, 1.853) | 0.309 | -3.705 (-10.074, 2.665) | 0.254 | 0.174 | NA |
| ALM | 40 | 28330 | 4.942 | 579.909 | -0.246 (-1.029, 0.536) | 0.537 | -3.276 (-7.231, 0.679) | 0.104 | 0.126 | NA |
| Male-specific |  |  |  |  |  |  |  |  |  |  |
| BMI | 38 | 152882 | 4.731 | 583.146 | 0.443 (0.163, 0.724) | 0.002 | 0.755 (-0.650, 2.159) | 0.292 | 0.657 |  |
| HC | 37 | 93480 | 4.522 | 571.095 | 0.228 (-0.102, 0.557) | 0.176 | 2.204 (0.492, 3.915) | 0.012 | 0.021 |  |
| WC | 35 | 93480 | 4.299 | 572.741 | 0.526 (0.238, 0.813) | <0.001 | 1.335 (-0.241, 2.911) | 0.097 | 0.306 |  |
| WHR | 38 | 93480 | 4.775 | 588.810 | 0.598 (0.300, 0.896) | <0.001 | -1.020 (-2.522, 0.482) | 0.183 | 0.032 |  |
| BMI-adjusted HC | 38 | 93480 | 4.723 | 582.123 | -0.368 (-0.673, -0.063) | 0.018 | 0.693 (-0.879, 2.266) | 0.388 | 0.178 |  |
| BMI-adjusted WC | 40 | 93480 | 5.133 | 603.571 | 0.079 (-0.156, 0.314) | 0.508 | 0.784 (-0.402, 1.969) | 0.195 | 0.235 |  |
| BMI-adjusted WHR | 40 | 93480 | 5.133 | 603.571 | 0.321 (0.094, 0.548) | 0.006 | -0.346 (-1.503, 0.811) | 0.557 | 0.249 |  |
| BF percentage | 41 | 44429 | 5.207 | 597.795 | 0.622 (0.268, 0.976) | 0.001 | -0.406 (-2.135, 1.323) | 0.645 | 0.234 |  |
| Female-specific |  |  |  |  |  |  |  |  |  |  |
| BMI | 36 | 171963 | 4.452 | 577.461 | 0.594 (0.333, 0.855) | <0.001 | 0.361 (-1.034, 1.755) | 0.612 | 0.739 |  |

|  |  |  |  |  |  |  |  |  |  |
| --- | --- | --- | --- | --- | --- | --- | --- | --- | --- |
| HC | 37 | 116742 | 4.669 | 590.581 | 0.437 (0.135, 0.739) | 0.005 | 0.710 (-0.926, 2.345) | 0.395 | 0.740 |
| WC | 33 | 116742 | 4.249 | 600.039 | 0.464 (0.170, 0.758) | 0.002 | -0.053 (-1.592, 1.485) | 0.946 | 0.502 |
| WHR | 37 | 116742 | 4.602 | 581.688 | 0.337 (0.039, 0.636) | 0.027 | 0.471 (-1.184, 2.125) | 0.577 | 0.872 |
| BMI-adjusted HC | 38 | 116742 | 4.822 | 594.864 | -0.028 (-0.240, 0.184) | 0.796 | 0.598 (-0.501, 1.697) | 0.286 | 0.256 |
| BMI-adjusted WC | 40 | 116742 | 5.212 | 613.340 | 0.135 (-0.067, 0.336) | 0.190 | 0.430 (-0.594, 1.453) | 0.411 | 0.564 |
| BMI-adjusted WHR | 38 | 116742 | 4.659 | 573.819 | 0.170 (-0.052, 0.391) | 0.133 | 0.702 (-0.534, 1.938) | 0.266 | 0.391 |
| BF percentage | 42 | 45525 | 5.319 | 596.862 | 0.334 (0.007, 0.662) | 0.045 | 0.713 (-0.908, 2.334) | 0.388 | 0.640 |

BMI: body mass index. WC: waist circumference. HC: hip circumference. WHR: waist-to-hip ratio. BF: body fat. ALM: appendicular lean mass. WLM: whole body lean mass. SNP: single nucleotide polymorphism. $R^2(\%)$ : the percentage of exposure variance explained by SNPs. 95% CI: 95% confidence interval. NA: not applicable.

In women, UNa increased BMI (0.571 (0.295 to 0.848); Figure 1(C)), BMI-adjusted WHR (0.249 (0.027 to 0.472); Figure 2(C)) and BF percentage (0.348 (0.027 to 0.669); Figure 3(C)). UNa was associated with HC and WC, but the associations became insignificant after adjustment for BMI. No pleiotropy was detected. (Table 1)

## Multivariable MR

The results of multivariable MR analyses were largely consistent with the univariable analyses. Overall, UNa was associated with increased BMI and BF percentage in both men and women. Higher UNa affected male body shape by increasing BMI-adjusted WHR and decreasing BMI-adjusted HC. UNa seemed to have little effects on female body shape after adjusted for BMI and eGFR. Formal heterogeneity test showed significant difference between men and women on BMI-adjusted HC, but not other outcomes. (**Table** 2) The eGFR-adjusted effect of BMI on UNa was 0.043 (0.023 to 0.063) with no pleiotropy (p = 0.149), indicating a bidirectional association between BMI and UNa.

In sensitivity analysis including the SNPs that were identified as outliers by MR-PRESSO outlier tests, both the univariable and multivariable MR showed results consistent with primary results. (**Supplement table 2** and **supplementary table 3**) Removing the eGFR-associated SNP rs1260326 also generated concordant results. (**Supplementary table 4**)

The scatter plots showed a potential outlier SNP rs4803378, located in gene *CTC-490E21.10* and associated with urine creatinine according to PhenoScanner v2 database, the *post hoc* sensitivity analyses by removing it showed similar results to the primary results. (**Supplementary table 5**)

## Discussion

In this MR study, salt intake, measured with UNa, was causally associated with increased BMI and BF percentage. Higher salt intake changed body shape towards central obesity in men, by reducing BMI-adjusted HC and increasing BMI-adjusted WHR, but probably not in women. Additionally, salt intake and BMI were bidirectionally positively related.

These results were consistent with previous findings. Moosavian *et al*.(18) found in a meta-analysis that both dietary salt intake and UNa were positively associated with BMI. This study computed the effect measure as the mean difference of BMI between highest and lowest categories of exposure, which introduced heterogeneity due to various cutoff values used and made it difficult to directly compare the effect size with other studies. Zhou *et al*.(19) found that for each 1gram per day (g/d) increase in salt intake, BMI increased by 0.42 and 0.52 kg/m^2^, overweight/obesity risk increased by 29% and 24% in UK and US population, respectively, after adjustment for total energy intake and physical activity..

Salt intake showed an effect on body composition by increasing BF percentage, which is consistent with previous findings. An increase of 1g/d salt intake was associated with an increase of 0.91kg BF mass in UK population (21), and of 0.79kg BF mass and 0.44% BF percentage in US population (20). Oh *et al*. (43) observed this positive association but only in people under 65 years old. Zhu *et al*. (44) found salt intake was associated with increased BF mass, BF percentage, and subcutaneous abdominal adipose tissue. Interestingly, a randomized trial (45) found that low-salt diet significantly decreased body weight and extracellular water, but reductions in BF mass and lean mass were insignificant. However, this trial was limited by small sample size (84 obese people) and short follow-up period (2 months), which might make it underpowered to detect significant reductions. Salt intake has an instant effect on water retention, but its effect on fat accumulation may take longer.

We found null effect of salt intake on lean mass, neither WLM nor ALM. This remains uncertain. Previously, Ma *et al*. (21) showed that 1 g/d increase in salt intake was associated with significant WLM increase by 0.32kg, but 1 g/2000kcal increase in salt intake density insignificantly reduced WLM by 0.007kg. Zhang *et al*. (20) found that salt intake significantly increased WLM by 0.53kg, but the effect became insignificant when using salt intake density (g/kcal) to adjust for energy intake. It seems that there are two possible effects of salt intake on body composition: (1) increase both fat and lean mass, but fat mass increases at a higher rate; (2) increase fat mass solely. The first explanation was supported by the findings that salt intake was associated with BF mass increase more than lean mass in both UK (0.91kg versus 0.32kg for fat and lean mass increase, respectively) (21) and US populations (0.79kg versus 0.53kg) (20). The second explanation was supported by the the null effect of salt intake on lean mass after adjustment for energy intake (by using salt intake density). (20,21) Our finding may also support the second explanation. Nevertheless, more research is warranted on this topic.

There are several potential explanations for how salt intake affects body weight and composition. It has been well recognized that excessive salt intake increases thirst and fluid consumption, thus increasing extracellular water while keeping urine volume almost unchanged. (46) Higher dietary salt intake also increases sweetened-beverage intake (47), food appetite and calorie intake (18,48), which all contribute to weight gain. In addition, high sodium intake has independent biological effects on fat accumulation. Lanaspa *et al*. (49) demonstrated that high salt intake activated the aldose reductase-fructokinase pathway in liver and hypothalamus, causing endogenous fructose production and the development of leptin resistance and hyperphagia, which finally cause obesity. Fonseca-Alaniz *et al*. (50) also observed that the high salt-induced adiposity in rats was characterized by high plasma leptin concentrations and adipocyte hypertrophy, which might be mediated by lipogenic capacity of white adipose mass. Lee *et al*. (51) revealed that high salt increased the expression of adipogenic and lipogenic genes (such as *PPAR-γ, SREBP1c, ACC* and *C/EBPα*), but decreased lipolysis gene expression (such as *AMPK*). However, its effect on lean mass requires more replications and mechanism research.

We found that salt intake had an impact on body shape. Body shapes more predictive of obesity-related mortality and health outcomes (52,53). Overall, salt intake increased both BMI-adjusted and unadjusted WHR, leading to central obesity, which is in line with previous findings (17,18,20,44). However, we observed an effect modification caused by sex. Salt intake increased WHR in both sexes, but a significant increase in BMI-adjusted WHR was seen in men only. Nam *et al*. (17) also found a similar sex difference. This may suggest that salt intake affects body shape independent of BMI in men, but its effect on female body shape is, at least partly, mediated by BMI.

This sexual dimorphism of body fat accumulation and distribution may be related to sex hormones (26,54), sex chromosome (55) and sex-specific autosomal genetic heritability (27,28,56,57). Estrogens favor fat accumulation in subcutaneous depot in women. Estrogens and their receptor regulation also enhance the expandability of adipose cells in subcutaneous depot and inhibits it in visceral depots. (26) Elevated free androgens have also been shown to be related to increased abdominal visceral fat accumulation and increased WC. (58) The Four Core Genotypes mouse model, has shown that X chromosome dosage is a risk factor for fat accumulation, but the presence of Y chromosome is not (56). Other studies have also shown that that adipocytes from different subcutaneous depots (abdominal versus gluteal) are developmentally distinct in a sex-dependent manner (59,60). The GWA study of BMI-adjusted WHR showed a higher heritability in women than in men (2.4% versus 0.8%) (32). This indicates that male body shape is more likely to be influenced by environmental factors (56), such as salt intake. However, mechanistic studies are needed to elucidate this issue.

In this MR study, we observed that high BMI was causally linked to higher UNa. High UNa could be resulted from two possible mechanisms: obesity-related glomerulopathy and high salt intake. Obesity is known to increase risk of glomerulopathy, characterized with altered renal hemodynamics, increased eGFR and tubular sodium reabsorption (61); however, the overall effect of glomerulopathy on UNa remains unclear. On the other hand, in our study, the association between BMI and UNa remained significant after adjustment for renal function, therefore, we believe that salt intake has independent mediation effect on the association between BMI and UNa, that is, BMI increases UNa by increasing salt intake, independent of renal function. Li *et al*. (62) found that overweight/obese people had lower salt sensitivity and higher salt preference, and consumed 2.3g/d more dietary salt than normal-weight people

The bidirectional association between obesity and salt intake indicates the formation of a vicious cycle, in which people with higher BMI are prone to consume more salt, which then further increases their BMI. Considering that high salt intake and obesity, both prevalent worldwide, are shared risk factors for many chronic diseases and that they also have their own adverse health consequences (63), urgent interventions are required to break this cycle in high-risk individuals.

Salt reduction and weight reduction programs have been widely implemented in various settings, but separately. Salt reduction programs do not consider weight change as an outcome, while weight reduction programs pay little emphasis on salt reduction measures. We are not advocating weight control by salt reduction alone. Instead, our findings provide evidence to combine salt reduction and weight reduction measures, to facilitate people to acquire a healthier lifestyle and achieve higher health benefits. Although the actual effect size of salt reduction on weight reduction has not been precisely estimated yet, which warrants future well-designed randomized trials, salt reduction *per se* costs less and brings additional health benefits. (22,43,64)

There are some limitations. First, we did not include all identified UNa-associated SNPs because unmatched data and pleiotropy issues; however, our results are consistent in primary analyses and sensitivity analyses, and with previous studies. Second, more accurate measurement of UNa is via 24-hour urine sample collection, but the UK Biobank uses spot urine sample. The 24-hour urine collection method is more expensive, time-consuming, labor-intensive and difficult for an epidemiological study as large as UK Biobank with half a million participants. Previous studies showed that spot UNa is an acceptable surrogate for 24-hour UNa and dietary salt intake, and has been commonly employed elsewhere (25,30). Third, there might still be residual pleiotropy, although we have removed SNPs that were significantly associated with outcomes and that were identified as outliers by the MR-PRESSO outlier test. MR-Egger intercept test showed pleiotropy for a few secondary outcomes (e.g., HC, WC) only. Fourth, the effect size estimated in this study was not directly comparable to other studies in which the effect was measured at natural units of outcome by 1 g/d salt intake increase. Although several equations have been proposed to estimate salt intake in g/d from 24h urine sodium secretion in g/d or spot urine sodium concentration in mmol/L (25), the UNa GWA study log-transformed the original spot UNa concentration, which makes back-transformation of beta estimate to natural unit impossible. Due to the same reason, formal power calculations were not performed as we planned, which was another limitation, but our study has employed strong instrumental variables and so far the largest GWA studies available. Lastly, we only included European-ancestry population, thus generalizing the results to other ethnicities requires cautions.

## Conclusion

Salt intake increases BMI and BF percentage, and changes male body shape towards central obesity by increasing BMI-adjusted WHR. Given the bidirectional association between salt intake and obesity, salt reduction measures and weight reduction measures should be implemented together to achieve more health benefits.

## Data Availability

Data are available upon request.

## Ethical approval

Not required.

## Data sharing

Data described in the manuscript, code book and analytic code will be made available upon request.

**Abbreviations**
ALM: appendicular lean mass
BF: body fat
BMI: body mass index
eGFR: estimated glomerular filtration rate
GWA: genome-wide association
HC: hip circumference
MR: Mendelian randomization
PRESSO test: Pleiotropy RESidual Sum and Outlier test
SNP: single nucleotide polymorphism
UNa: urinary sodium secretion
WC: waist circumference
WHR: waist-to-hip ratio
WLM: whole body lean mass

## Reference

1. Tremmel M, Gerdtham UG, Nilsson PM, Saha S. Economic burden of obesity: a systematic literature review. Int J Environ Res Public Health. 2017;14.

2. Seidell JC, Halberstadt J. The global burden of obesity and the challenges of prevention. Ann Nutr Metab. 2015;66:7–12.

3. Arroyo-Johnson C, Mincey KD. Obesity epidemiology worldwide. Gastroenterol Clin North Am. 2016;45:571–9.

4. Apovian CM. Obesity: definition, comorbidities, causes, and burden. Am J Manag Care. 2016;22:s176–85.

5. Lee EY, Yoon KH. Epidemic obesity in children and adolescents: risk factors and prevention. Front Med. 2018;12:658–66.

6. Ortega FB, Lavie CJ, Blair SN. Obesity and cardiovascular disease. Circ Res. 2016;118:1752–70.

7. Malone JI, Hansen BC. Does obesity cause type 2 diabetes mellitus (T2DM)? Or is it the opposite? Pediatr Diabetes. 2019;20:5–9.

8. Wei J, Liu X, Xue H, Wang Y, Shi Z. Comparisons of visceral adiposity index, body shape index, body mass index and waist circumference and their associations with diabetes mellitus in adults. Nutrients. 2019; 11.

9. Avgerinos KI, Spyrou N, Mantzoros CS, Dalamaga M. Obesity and cancer risk: Emerging biological mechanisms and perspectives. Metabolism. 2019;92:121–35.

10. De Pergola G, Silvestris F. Obesity as a major risk factor for cancer. J Obes. 2013;2013:291546.

11. Gunter MJ, Riboli E. Obesity and gastrointestinal cancers - where do we go from here? Nat Rev Gastroenterol Hepatol. 2018;15:651–2.

12. Chen G-C, Arthur R, Iyengar NM, Kamensky V, Xue X, Wassertheil-Smoller S, Allison MA, Shadyab AH, Wild RA, Sun Y, et al. Association between regional body fat and cardiovascular disease risk among postmenopausal women with normal body mass index. Eur Heart J. 2019;40:2849–55.

13. Huxley R, Mendis S, Zheleznyakov E, Reddy S, Chan J. Body mass index, waist circumference and waist:hip ratio as predictors of cardiovascular risk--a review of the literature. Eur J Clin Nutr. 2010;64:16–22.

14. Rust P, Ekmekcioglu C. Impact of salt intake on the pathogenesis and treatment of hypertension. Adv Exp Med Biol. 2017;956:61–84.

15. Meneton P, Jeunemaitre X, de Wardener HE, MacGregor GA. Links between dietary salt intake, renal salt handling, blood pressure, and cardiovascular diseases. Physiol Rev. 2005;85:679–715.

16. Horikawa C, Sone H. Dietary salt intake and diabetes complications in patients with diabetes: An overview. J Gen Fam Med. 2017;18:16–20.

17. Nam GE, Kim SM, Choi MK, Heo YR, Hyun TS, Lyu ES, Oh SY, Park HR, Ro HK, Han K, et al. Association between 24-h urinary sodium excretion and obesity in Korean adults: A multicenter study. Nutrition. 2017;41:113–9.

18. Moosavian SP, Haghighatdoost F, Surkan PJ, Azadbakht L. Salt and obesity: a systematic review and meta-analysis of observational studies. Int J Food Sci Nutr. 2017;68:265–77.

19. Zhou L, Stamler J, Chan Q, Van Horn L, Daviglus ML, Dyer AR, Miura K, Okuda N, Wu Y, Ueshima H, et al. Salt intake and prevalence of overweight/obesity in Japan, China, the United Kingdom, and the United States: the INTERMAP Study. Am J Clin Nutr. 2019;110:34–40.

20. Zhang X, Wang J, Li J, Yu Y, Song Y. A positive association between dietary sodium intake and obesity and central obesity: results from the National Health and Nutrition Examination Survey 1999–2006. Nutr Res. 2018;55:33–44.

21. Ma Y, He FJ, MacGregor GA. High salt intake: independent risk factor for obesity? Hypertension. 2015;66:843–9.

22. He FJ, Li J, MacGregor GA. Effect of longer term modest salt reduction on blood pressure: Cochrane systematic review and meta-analysis of randomised trials. BMJ. 2013;346:f1325-f1325.

23. Hyseni L, Elliot-Green A, Lloyd-Williams F, Kypridemos C, O’Flaherty M, McGill R, Orton L, Bromley H, Cappuccio FP, Capewell S. Systematic review of dietary salt reduction policies: Evidence for an effectiveness hierarchy? PloS One. 2017;12:e0177535.

24. Emdin CA, Khera AV, Kathiresan S. Mendelian randomization. JAMA. 2017;318:1925–6.

25. Huang L, Crino M, Wu JHY, Woodward M, Barzi F, Land M-A, McLean R, Webster J, Enkhtungalag B, Neal B. Mean population salt intake estimated from 24-h urine samples and spot urine samples: a systematic review and meta-analysis. Int J Epidemiol. 2016;45:239–50.

26. Palmer BF, Clegg DJ. The sexual dimorphism of obesity. Mol Cell Endocrinol. 2015;402:113–9.

27. Rask-Andersen M, Karlsson T, Ek WE, Johansson A. Genome-wide association study of body fat distribution identifies adiposity loci and sex-specific genetic effects. Nat Commun. 2019;10:339.

28. Winkler TW, Justice AE, Graff M, Barata L, Feitosa MF, Chu S, Czajkowski J, Esko T, Fall T, Kilpelainen TO, et al. The influence of age and sex on genetic associations with adult body size and shape: A large-scale genome-wide interaction study. PLoS Genet. 2015;11:e1005378.

29. Pazoki R, Evangelou E, Mosen-Ansorena D, Pinto RC, Karaman I, Blakeley P, Gill D, Zuber V, Elliott P, Tzoulaki I, et al. GWAS for urinary sodium and potassium excretion highlights pathways shared with cardiovascular traits. Nat Commun. 2019;10:3653.

30. Welsh CE, Welsh P, Jhund P, Delles C, Celis-Morales C, Lewsey JD, Gray S, Lyall D, Iliodromiti S, Gill JMR, et al. Urinary sodium excretion, blood pressure, and risk of future cardiovascular disease and mortality in subjects without prior cardiovascular disease. Hypertens Dallas Tex 1979. 2019;73:1202–9.

31. Locke AE, Kahali B, Berndt SI, Justice AE, Pers TH, Day FR, Powell C, Vedantam S, Buchkovich ML, Yang J, et al. Genetic studies of body mass index yield new insights for obesity biology. Nature. 2015;518:197–206.

32. Shungin D, Winkler TW, Croteau-Chonka DC, Ferreira T, Locke AE, Magi R, Strawbridge RJ, Pers TH, Fischer K, Justice AE, et al. New genetic loci link adipose and insulin biology to body fat distribution. Nature. 2015;518:187–96.

33. Zillikens MC, Demissie S, Hsu YH, Yerges-Armstrong LM, Chou WC, Stolk L, Livshits G, Broer L, Johnson T, Koller DL, et al. Large meta-analysis of genome-wide association studies identifies five loci for lean body mass. Nat Commun. 2017;8:80.

34. Lu Y, Day FR, Gustafsson S, Buchkovich ML, Na J, Bataille V, Cousminer DL, Dastani Z, Drong AW, Esko T, et al. New loci for body fat percentage reveal link between adiposity and cardiometabolic disease risk. Nat Commun. 2016;7:10495.

35. Nomura K, Asayama K, Jacobs L, Thijs L, Staessen JA. Renal function in relation to sodium intake: a quantitative review of the literature. Kidney Int. 2017;92:67–78.

36. Jamshidi P, Najafi F, Mostafaei S, Shakiba E, Pasdar Y, Hamzeh B, Moradinazar M. Investigating associated factors with glomerular filtration rate: structural equation modeling. BMC Nephrol. 2020;21:30.

37. Whaley-Connell A, Sowers JR. Obesity and kidney disease: from population to basic science and the search for new therapeutic targets. Kidney Int. 2017;92:313–23.

38. Wuttke M, Li Y, Li M, Sieber KB, Feitosa MF, Gorski M, Tin A, Wang L, Chu AY, Hoppmann A, et al. A catalog of genetic loci associated with kidney function from analyses of a million individuals. Nat Genet. 2019;51:957–72.

39. Verbanck M, Chen CY, Neale B, Do R. Detection of widespread horizontal pleiotropy in causal relationships inferred from Mendelian randomization between complex traits and diseases. Nat Genet. 2018;50:693–8.

40. Pierce BL, Ahsan H, Vanderweele TJ. Power and instrument strength requirements for Mendelian randomization studies using multiple genetic variants. Int J Epidemiol. 2011;40:740–52.

41. Hartwig FP, Davies NM, Hemani G, Davey Smith G. Two-sample Mendelian randomization: avoiding the downsides of a powerful, widely applicable but potentially fallible technique. Int J Epidemiol. 2016;45:1717–26.

42. Brion M-JA, Shakhbazov K, Visscher PM. Calculating statistical power in Mendelian randomization studies. Int J Epidemiol. 2013;42:1497–501.

43. Oh SW, Koo HS, Han KH, Han SY, Chin HJ. Associations of sodium intake with obesity, metabolic disorder, and albuminuria according to age. PloS One. 2017;12:e0188770.

44. Zhu H, Pollock NK, Kotak I, Gutin B, Wang X, Bhagatwala J, Parikh S, Harshfield GA, Dong Y. Dietary sodium, adiposity, and inflammation in healthy adolescents. Pediatrics. 2014;133:e635–642.

45. Kang HJ, Jun DW, Lee SM, Jang EC, Cho YK. Low salt and low calorie diet does not reduce more body fat than same calorie diet: a randomized controlled study. Oncotarget. 2018;9:8521–30.

46. Bankir L, Perucca J, Norsk P, Bouby N, Damgaard M. Relationship between sodium intake and water intake: the false and the true. Ann Nutr Metab. 2017;70 Suppl 1: 51–61.

47. Grimes CA, Riddell LJ, Campbell KJ, Nowson CA. Dietary salt intake, sugar-sweetened beverage consumption, and obesity risk. Pediatrics. 2013;131:14–21.

48. Bolhuis DP, Costanzo A, Newman LP, Keast RS. Salt promotes passive overconsumption of dietary fat in humans. J Nutr. 2016;146:838–45.

49. Lanaspa MA, Kuwabara M, Andres-Hernando A, Li N, Cicerchi C, Jensen T, Orlicky DJ, Roncal-Jimenez CA, Ishimoto T, Nakagawa T, et al. High salt intake causes leptin resistance and obesity in mice by stimulating endogenous fructose production and metabolism. Proc Natl Acad Sci U A. 2018;115:3138–43.

50. Fonseca-Alaniz MH, Brito LC, Borges-Silva CN, Takada J, Andreotti S, Lima FB. High dietary sodium intake increases white adipose tissue mass and plasma leptin in rats. Obes Silver Spring. 2007;15:2200–8.

51. Lee M, Sorn SR, Lee Y, Kang I. Salt induces adipogenesis/lipogenesis and inflammatory adipocytokines secretion in adipocytes. Int J Mol Sci. 2019;20(1):pii.

52. Fu J, Hofker M, Wijmenga C. Apple or pear: size and shape matter. Cell Metab. 2015;21:507–8.

53. Golzarri-Arroyo L, Mestre LM, Allison DB. What’s new in understanding the risk associated with body size and shape?: pears, apples, and olives on toothpicks. JAMA Netw Open. 2019;2:e197336.

54. Valencak TG, Osterrieder A, Schulz TJ. Sex matters: The effects of biological sex on adipose tissue biology and energy metabolism. Redox Biol. 2017;12:806–13.

55. Zore T, Palafox M, Reue K. Sex differences in obesity, lipid metabolism, and inflammation-A role for the sex chromosomes? Mol Metab. 2018;15:35–44.

56. Link JC, Reue K. Genetic basis for sex differences in obesity and lipid metabolism. Annu Rev Nutr. 2017;37:225–45.

57. Pulit SL, Karaderi T, Lindgren CM. Sexual dimorphisms in genetic loci linked to body fat distribution. Biosci Rep. 2017;37.

58. Blouin K, Boivin A, Tchernof A. Androgens and body fat distribution. J Steroid Biochem Mol Biol. 2008;108:272–80.

59. Fried SK, Lee M-J, Karastergiou K. Shaping fat distribution: New insights into the molecular determinants of depot- and sex-dependent adipose biology. Obes Silver Spring Md. 2015;23:1345–52.

60. Karastergiou K, Fried SK, Xie H, Lee M-J, Divoux A, Rosencrantz MA, Chang RJ, Smith SR. Distinct developmental signatures of human abdominal and gluteal subcutaneous adipose tissue depots. J Clin Endocrinol Metab. 2013;98:362–71.

61. D’Agati VD, Chagnac A, de Vries APJ, Levi M, Porrini E, Herman-Edelstein M, Praga M. Obesity-related glomerulopathy: clinical and pathologic characteristics and pathogenesis. Nat Rev Nephrol. 2016;12:453–71.

62. Li Q, Jin R, Yu H, Lang H, Cui Y, Xiong S, Sun F, He C, Liu D, Jia H, et al. Enhancement of neural salty preference in obesity. Cell Physiol Biochem. 2017;43:1987–2000.

63. Cheung BMY, Li C. Diabetes and hypertension: is there a common metabolic pathway? Curr Atheroscler Rep. 2012;14:160–6.

64. Hoffmann IS, Cubeddu LX. Salt and the metabolic syndrome. Nutr Metab Cardiovasc Dis NMCD. 2009;19:123–8.

